# Biochemical fingerprinting of human scalp hair reveals endocannabinoid-related compounds as potential biomarker indicators of altered mitochondrial bioenergetics in immune cells from female patients with major depressive disorder

**DOI:** 10.64898/2026.06.15.26355692

**Authors:** Leonard Bondy, Karin de Punder, Juan Salinas-Manrique, Thomas Hennessy, Thomas Stoll, Michelle M. Hill, Detlef E. Dietrich, Alexander Karabatsiakis

**Affiliations:** University of Innsbruck, Department of Psychology, Clinical Psychology II, 6020 Innsbruck, Austria; AMEOS Clinic for Psychiatry and Psychotherapy Hildesheim, 31135 Hildesheim, Germany; Agilent Technologies, 679 Springvale Road, Mulgrave, VIC 3170, Australia; University of Queensland Diamantina Institute, The University of Queensland, Translational Research Institute, 37 Kent Street, Woolloongabba, QLD 4102, Australia

**Keywords:** Major Depressive Depression, Scalp hair, Mass spectrometry, Biochemical fingerprinting, Endocannabinoids, PBMC, Mitochondria, Bioenergetics, Biomarkers

## Abstract

Major depressive disorder (MDD) is a severe psychiatric disorder that affects more than 350 million people worldwide, yet its biomolecular mechanisms are incompletely understood, and clinically applicable markers remain elusive. To shed new light on the underlying pathophysiology of MDD across multiple research disciplines, we first used a biochemical fingerprinting approach with human hair (the first 3 cm cut from the scalp) to identify changes in the total set of detectable metabolites and lipids (metabolipidomics) using quadrupole time-of-flight mass spectrometry (qToF-MS). In this study, we focused on endocannabinoid (ECB)-related lipid compounds and identified 7 candidate markers that differed between depressed and non-depressed female participants. Two phosphatidylinositols, namely PI 24:0 and PI 37:4, showed dose-dependent associations with the severity of depressive symptoms. Finally, to bridge hair findings with previously reported results in blood, we tested associations between changes in identified ECB-related compounds and parameters of mitochondrial respiratory activity in peripheral blood mononuclear cells. We found 17 significant associations, with the strongest effects for the lipids PI 24:0, MGDG-O 16:3, PG 12:0, and PI 37:4. Our approach not only identified novel associations between endocannabinoid (ECB)-related lipid dysregulation and impaired mitochondrial energy metabolism in MDD but also revealed ECB-related lipids as a possible surrogate marker of impaired bioenergetic metabolism in MDD, at least in immune cells. More research is needed to replicate these findings, ideally by testing reversibility in longitudinal intervention studies and by including both sexes in larger cohorts.

## Introduction

Major depressive disorder (MDD) is a severe psychiatric disorder characterized by substantial impairments in psychosocial functioning and well-being (Christensen et al., 2020). The clinical presentation of MDD varies widely among individuals (Kennedy, 2008; Otte et al., 2016). Some predominantly exhibit sadness and hopelessness, while others may experience pronounced cognitive symptoms, such as difficulty concentrating or making decisions (Knight and Baune, 2018; Otte et al., 2016). Additionally, MDD can manifest with physical symptoms including sleep disturbances, changes in appetite or weight, altered pain perception, fatigue, or lack of energy (Fava et al., 2014; Vaccarino et al., 2008). According to the *World Health Organization* (WHO), depression has become the leading cause of disability worldwide, especially among those ages 15-44 (Global Burden of Disease Collaborative Network, 2021; The Lancet Global Health, 2020). The prevalence is considerable, with approximately 5-10% of men and 10-25% of women at risk of experiencing a depressive episode at some point in their lives (Gutiérrez-Rojas et al., 2020; Otte et al., 2016).

The etiology of MDD is complex, involving a multitude of psychological, social, and biological factors that shape an individual’s vulnerability or resilience to the disorder. One of the most consistent findings is the association between adverse childhood experiences (ACEs), such as neglect, abuse, or parental loss, and the later development of MDD (Antoniou et al., 2023; Chapman et al., 2004; Kuehl et al., 2022). Furthermore, cognitive vulnerabilities, including negative cognitive styles, rumination, maladaptive cognitive biases, and certain personality traits, are linked to increased risk of MDD (Chapman et al., 2004; Kuehl et al., 2022; Robinson, 2003). Etiological models that emphasize biological vulnerability include genetic predispositions and neurobiological mechanisms, particularly the roles of the monoamines serotonin and norepinephrine and of brain regions implicated in emotional processing, such as the amygdala and the prefrontal cortex (PFC) (Hymie Anisman, 2009; Lau et al., 2009; Lohoff, 2010; Raedler, 2011; Shadrina et al., 2018; Shea et al., 2005; Slavich and Irwin, 2014). Additionally, inflammation and altered levels of stress hormones related to dysregulation of the hypothalamus-pituitary-adrenal (HPA) axis are prominent findings in individuals with MDD, supporting the idea that altered stress and immune responses represent supplementary characteristics of the disorder (Ceruso et al., 2020; Hymie Anisman, 2009; Karabatsiakis et al., 2022a, 2014; Raedler, 2011; Shea et al., 2005; Slavich and Irwin, 2014). In line with this, associations between MDD and various metabolic conditions (e.g., obesity, dyslipidemia, coronary diseases, diabetes, high blood pressure) point to a link between molecular stress, dysregulation of fat or energy metabolism, and depressive symptomatology (Boeck et al., 2018a, 2018b; McIntyre et al., 2009; Pitsillou et al., 2020; Stapelberg et al., 2012; Stunkard et al., 2003; Vancampfort et al., 2014; Zhu et al., 2023). Thus, to promote resilience, a well-functioning stress response and a healthy metabolism may be as important as psychological and social factors.

Although many of the comorbid conditions associated with MDD can be measured, e.g., by assessing inflammatory or immune markers (i.e., cortisol, TNF-alpha, CRP, IL-6) or metabolic indicators (Kim et al., 2007; McIntyre, 2021; Young et al., 2014), there remains a lack of disease-specific biomarkers that can be applied to enhance treatment and prevention strategies for MDD. This is important because the chronicity of the disease burden negatively affects treatment response and remission stability (Kraus et al., 2019). However, a biological entity for diagnosis, as well as for assessing reversibility after clinical intervention, is missing. Over the last decade, the endocannabinoid system (ECS) has attracted promising attention in this regard. The ECS is a neuromodulatory, lipid-based signaling system distributed throughout the body that plays a critical role in regulating neuronal signaling and the molecular stress response (Bambico et al., 2007; Haj-Dahmane and Shen, 2011; Morena et al., 2016; Pandey et al., 2009; Turcotte et al., 2015). It is considered to play a central part in the establishment and maintenance of homeostasis and is assumed to function as a gatekeeper of the HPA axis, thus regulating the endocrine secretion of glucocorticoids such as cortisol (Boczek and Zylinska, 2021; Morena et al., 2016).

Three primary elements form the ECS: two main cannabinoid receptors (CB1 and CB2) that modulate neuronal signaling; endocannabinoids (ECBs) such as arachidonoyl ethanolamide (AEA) and 2-arachidonoyl glycerol (2-AG) that serve as ligands; and enzymes such as *fatty acid amid hydrolase* (FAAH) and *monoglyceride lipase* (MAGL) that catabolize them. CB1 is predominantly expressed in the central nervous system (CNS), particularly in the cortex, hippocampus, basal ganglia, and cerebellum, and is located presynaptically on the axon terminals of serotonergic, noradrenergic, and dopaminergic neurons. CB2 is mainly found in immune cells, the spleen, and the tonsils, and modulates immune cell migration and cytokine release upon activation (Battista et al., 2012; Kano et al., 2002; Silver, 2019). ECB signaling is thought to play a central role in regulating neural circuits associated with depressive symptomatology and stress (McLaughlin et al., 2011; Morena et al., 2016). This includes neuromodulatory effects on psychological features such as mood, anxiety, reward-seeking, pain, and appetite (Gray et al., 2015; Hillard and Liu, 2014; Huang et al., 2016; Mikics et al., 2009), as well as regulation of biological processes such as inflammatory and molecular stress responses (Gray et al., 2015; M. N. Hill et al., 2008; Morena et al., 2016; Wang et al., 2012). Many studies investigating these links in the context of MDD have found alterations in various ECB-related agents in patients with MDD (Bersani et al., 2021; Herranz-Herrer et al., 2020; M. Hill et al., 2008; Lazary et al., 2021). In line with this, most preclinical and clinical studies have shown that interventions that reduce ECB signaling can create or aggravate depressive symptoms, whereas promoting ECB activity shows antidepressant effects (Dong et al., 2020; Maccarrone et al., 2002; Malone et al., 2008; Rossi et al., 2010; Valverde and Torrens, 2012). While the main agents of the ECS have been extensively investigated in the context of mental disorders, little is known about the molecular structures and downstream pathways involved in ECB signaling, as well as their interactions with other biological systems involved in the physiological stress response. In line with this, the cellular energy supply via mitochondrial respiration has become a major focus of basic and clinical research on mental health and disease.

Mitochondria, often called the “powerhouses of the cell,” use molecular oxygen (O2) and high-energy carbohydrates to synthesize adenosine triphosphate (ATP) through oxidative phosphorylation (OXPHOS). ATP is the currency of biochemical energy and enables a cell to perform work (Picard and McEwen, 2018). In recent years, mitochondrial bioenergetics has emerged as a highly relevant research field for mental diseases, and disturbances in mitochondrial energy production are considered a key underlying mechanism of pathophysiological conditions associated with MDD (Allen et al., 2018; Boeck et al., 2018b; Karabatsiakis et al., 2020, 2014; Picard et al., 2014; Picard and McEwen, 2018).

Multiple parameters of mitochondrial activity, such as basal oxygen consumption, respiration related to ATP production, spare respiratory capacity, and mitochondrial coupling efficacy, were reported to be reduced in peripheral blood mononuclear cells (PBMCs) from individuals diagnosed with MDD (Boeck et al., 2018b; Karabatsiakis et al., 2020, 2014; Karabatsiakis and Schönfeldt-Lecuona, 2020). In addition, parameters of mitochondrial biogenesis, such as mitochondrial density measured as mtDNA copy number and the activity of the enzyme *citrate synthase*, were also affected (Allen et al., 2018; Boeck et al., 2018b; Karabatsiakis et al., 2020; Karabatsiakis and Schönfeldt-Lecuona, 2020; Klinedinst and Regenold, 2015; Lindqvist et al., 2016; Picard and Turnbull, 2013; Tyrka et al., 2016). These observations led to the idea that a lack of bioenergetic supply might explain the lack of drive and energy and/or the anhedonia symptoms of MDD. Furthermore, it is proposed that monoamine shortages in depressed patients might alternatively be explained by bioenergetic disturbances, since monoamine synthesis, like almost all processes in the organism, requires ATP (Allen et al., 2018; Hroudova and Fisar, 2010; Nahon et al., 2005; Picard and McEwen, 2018). Mitochondrial involvement in monoamine metabolism extends beyond energy production, as mitochondria are also involved in the production and catabolism of monoamines through several direct and indirect pathways (Graves et al., 2020; Nagatsu et al., 2019; Nahon et al., 2005; Tanaka et al., 2022).

Finally, mitochondria might play a crucial role in the emergence of inflammation in patients with MDD. Reactive oxygen species (ROS) are central drivers of pro-inflammatory signaling in the body and arise mainly within mitochondria, especially under stress and when mitochondrial respiration is impaired (Picard et al., 2014). In sum, the bioenergetic concept of mental diseases suggests that MDD is not primarily a disease of the brain, but rather that the brain is the most vulnerable organ to inflammation and to disturbances in energy supply (Karabatsiakis and Schönfeldt-Lecuona, 2020; Picard et al., 2014). To bridge these two fields, the ECS can interact with mitochondria in multiple ways. Because disturbances in mitochondrial respiration can result from endocrine cortisol excess (Klinedinst and Regenold, 2015; Picard et al., 2014), the ECS’s regulatory function on the HPA axis can affect both energy production and ROS generation, leading to inflammation (Paloczi et al., 2018; Pandey et al., 2009; Raja et al., 2020; Turcotte et al., 2015). Changes in ECB activity alter phospholipid demand and might therefore change the fluidity of the dynamic membrane, facilitating or complicating the uptake of molecules such as glucose, lipids, or cortisol into the cell. Because mitochondrial function depends on these molecules entering the cell, the interaction between ECB activity and the dynamic composition of the lipid cell membrane may be critical for efficient mitochondrial respiration. In addition, several mechanisms by which ECB activity can directly modulate mitochondrial functions have been proposed, including ECB-regulated levels of calcium, nitric oxide, and ceramide, as well as ECB-induced changes in the lipid composition of the cell membrane (Nunn et al., 2012). Recent interest has focused on CBR1s located within the outer mitochondrial membrane (Maya-López et al., 2021; Skupio et al., 2023). It is suggested that their activation facilitates cellular respiration, thereby promoting homeostasis by modulating energy production, apoptosis, and the cellular stress response (Bénard et al., 2012). In contrast to previous studies that primarily analyzed the ECB-related lipid composition of serum samples, the current study uses data from quadrupole time-of-flight mass spectrometry (qTOF-MS)-derived hair analyses to measure potential differences in hair ECB and ECB-related lipid levels between individuals diagnosed with MDD and non-depressed controls. Analyzing hair samples has certain advantages over biofluids (serum, plasma, saliva, or urine). Stress-related processes vary strongly with the time of day (circadian rhythm) and with factors such as recent physical activity, diet, and alcohol and nicotine consumption, making it difficult to detect consistent alterations (Stalder and Kirschbaum, 2012). By contrast, hair analysis is more robust to these factors and is considered a stable method for the retrospective assessment of cumulative metabolic processes, particularly relevant for ECBs (Krumbholz et al., 2013). Since hair growth in adults is estimated at about 1 cm per month, analyzing 3 cm provides information on ECB activity over the past 3 months (Wennig, 2000).

The first aim of the study was to perform biochemical fingerprinting of hair samples from females with and without MDD (data not shown). Subsequently, a range of ECB-related lipid molecules was targeted to assess their association with changes in mitochondrial energy metabolism, as previously reported in PBMCs from individuals with depression (Karabatsiakis et al., 2014). The molecules studied comprised various glycerophospholipid (GP) species, including phosphatidylcholines (PC), phosphatidylethanolamines (PE), and phosphatidylinositols (PI). In MDD research, analyses of genomics, metabolomics, or proteomics profiles are emerging as promising tools that can distinguish samples from depressed individuals from those of non-depressed controls, with area under the curve (AUC) rates of 82-96% (Liu et al., 2016; Pan et al., 2018; Zheng et al., 2012). So far, lipidomics analyses in human subjects present a somewhat inconsistent picture. One study found significantly elevated total levels of LPS, PC, PE, PI, and TAG species in depressed subjects (Liu et al., 2016), while another found reduced levels of PC O 36:4 to be the most robust finding associated with depressive symptoms (Demirkan et al., 2013). A proposed combinatorial lipid marker that achieved high AUC values (0.855–0.931) included the following five lipids: LPE 20:4, PC 34:1, PI 40:4, Sphingomyelin (SM) 39:1, and TAG 44:2 (Liu et al., 2016). In another study, increased TAG 44:0 and TAG 54:8, together with LPA 16:1, were the strongest discriminators, whereas PC and PE species showed no association with MDD (Kim et al., 2018). In a previous study using serum from individuals with post-traumatic stress disorder (PTSD), the ECB-related molecules PE(17:1(9Z)/18:0) and PEA showed the highest discriminative ability in segregating samples from patients with PTSD from trauma-exposed individuals without PTSD (Karabatsiakis et al., 2015). PEA is pivotal in the ECS and potentiates the effects of AEA. To explore possible associations among MDD, bioenergetic function in PBMC, and lipid compounds in hair samples, data from two groups, each with n=22 female participants aged 50-69, were analyzed. Endocannabinoid-related lipids were first identified and then tested for group differences, associations with depressive symptom severity (self-report vs. external assessment via interview), and associations with parameters of mitochondrial bioenergetic function measured in PBMC, as reported previously (Karabatsiakis et al., 2014).

## Methods

### Recruitment and clinical characterization of participants

Patients diagnosed with MDD were clinical inpatients receiving treatment at the AMEOS Clinic in Hildesheim, Germany. Control group participants were recruited via public advertisement from the same location. After providing written informed consent to a study protocol approved by the Local Ethics Board of the Hanover Medical School (Germany), the clinical severity of depressive symptoms was assessed by self-report in both groups using the German version of the *Beck Depression Inventory*-II (Hautzinger et al., 2006), followed by the German version of the *Montgomery-Asberg Depression Rating Scale* (MADRS) administered during an interview. For the biological components of the study, all participants were asked to donate a small volume of venous blood (35 ml, non-fasting) for isolation of peripheral blood mononuclear cells (PBMC) and to provide a hair sample. For a detailed description of the criteria for participation, procedures, and methods, see previously reported data (Boeck et al., 2018a).

### Collection, pre-analytical processing, and preparation of hair samples

Using sterile tools and gloves, hair samples at least 40 mm in length were collected from the posterior vertex of the skull, as previously reported (Boeck et al., 2018a; Hu et al., 2009; Karabatsiakis et al., 2022b, 2014), to avoid external contamination during collection. Hair strands were washed 3 × 3 min with HPLC-certified isopropanol (Sigma Aldrich, USA) and then lyophilized to complete dryness using a freeze dryer (Labconco, USA). Next, hair samples were cut into 3-cm segments, representing, on average, the past 3 months before sampling. Hair segments were then pulverized in a ball mill (MP Biomedicals, Germany) for 3 × 5 min at 30 Hz, and metabolites and lipids were extracted using methanol (Sigma Aldrich, USA) as previously reported (Karabatsiakis et al., 2022b, 2022a). Extracts were lyophilized again and shipped on dry ice to the Translational Research Institute (TRI) in Queensland, Australia. All subsequent steps were performed in a cold room (4 °C) due to the relatively high volatility of the solvents. Reconstitution of the lyophilized samples was performed with 80 µl of HPLC-certified methanol (VWR International), followed by short-term sonication for 20 sec, pulse vortexing, and centrifugation for 15 min at 15,000 g in an Eppendorf centrifuge set to 4 °C. After centrifugation, 35 µl of the clear supernatant was transferred into an MS glass vial (Agilent Technologies, USA), and an additional 4 µl per sample was used to generate a pooled quality control (QC) sample.

### Mass spectrometric analysis of hair extracts

The LC/MS platform consisted of a 1290 Infinity 8 II UHPLC coupled to a 6550 iFunnel Q-TOF mass spectrometer via a Dual AJS ESI source 9 (Agilent, Santa Clara, USA). Metabolite and lipid separation was performed on a Poroshell 120 EC-C18 column (2.7 µm, 120 Å, 2.1 × 100 mm, pn 695775-902, Agilent). The column was connected to a 2.1 x 5 mm guard column containing the same resin. For detailed descriptions of solvents, technical specifications, methods, procedures, and data processing, see the previous reports (Koenig et al., 2018).

### Statistical analysis

Data analysis was conducted using IBM® SPSS® Statistics 26 (IBM Corp.). The critical *p*-value for statistical significance was set at 0.05, corresponding to a 95% confidence level. For ECB-related molecules in hair, mass spectrometry-detected candidates were compared with the *Lipidmaps* database to identify those involved in the ECS. For group comparisons and dose-response analyses, only lipid compounds with data available for more than 50% of participants in each group were included. This criterion was met by 15 compounds. Shapiro-Wilk tests were conducted, and nonparametric analyses were used when residuals did not follow a normal distribution. To assess potential group differences in sociodemographic, clinical, and biological variables, Chi-squared tests were used for categorical variables, and two-tailed t-tests (parametric) or Mann-Whitney U-tests (non-parametric) were used for continuous variables. To analyze inter-correlations among lipid compounds and between lipid compounds and measures of mitochondrial respiration, Pearson correlations were calculated. Given the linear relationships and the overall distribution of the data, parametric tests were considered appropriate. Additionally, Kendall’s tau correlations were calculated to assess associations between measures of mitochondrial respiration and the severity of depressive symptoms, as measured by BDI (self-report) and MADRS sum scores (interview). Nonparametric group comparisons between the control and MDD groups were conducted for each of the 15 lipid compound candidates. To avoid overestimating the significance of the p-value due to alpha error accumulation in repeated hypothesis testing and to reduce the probability of committing type 1 errors, the p-values from the group comparisons were adjusted using the Bonferroni-Holm correction. Finally, for lipid compounds that showed significant between-group differences, separate linear regression models were fit using both BDI and MADRS sum scores to explore potential dose-response effects.

## Results

Sociodemographic and clinical variables are summarized in Table 1. A list of all lipids detected by the mass spectrometer, along with an intercorrelation matrix among those lipids, is also provided in the appendix (Appendices A and B).

**Table 1.** Sociodemographic, clinical, and lifestyle-related information on the GerontoTel study cohort.

|  | Controls | MDD |  |  |  |
| --- | --- | --- | --- | --- | --- |
| | (n = 22) | (n = 22) | W/$\chi^2a$ | df | p |
| Age (mean ± SD; years) | 57.6 ± 5.6 | 58.7 ± 6.1 | 219.0 | | 0.59 |
| BMI (mean ± SD; kg/m<sup>2</sup>) | 24.4 ± 3.0 | 28.5 ± 7.2 | 155.0 | | **0.04\*** |
| Smoking status (yes, n [%]) | 4 (18%) | 12 (55%) | 6.29 | 1 | **0.03\*** |
| Physical activity (yes, n [%]) | 19 (86%) | 14 (64%) | 3.03 | 1 | 0.82 |
| BDI sum score (mean ± SD) | 2.3 ± 2.3 | 24.0 ± 10.4 | 0.50 | | **< 0.001\*\*** |
| MADRS sum score (mean ± SD) | 2.0 ± 2.3 | 23.4 ± 8.9 | 0.50 | | **< 0.001\*\*** |
| *Medication intake* |  |  |  |  |  |
| Antidepressants (yes, N [%]) | - | 17 (77%) |  |  |  |
| Antipsychotics (yes, N [%]) | - | 8 (36%) |  |  |  |

\* $p \leq 0.5$ , \*\* $p < 0.01$
*Notes:* MDD = Major Depressive Disorder; BMI = Body Mass Index; BDI = Beck's Depression Inventory; MADRS = Montgomery-Asberg Depression Rating Scale; W = Mann-Whitney-U Test; $\chi^2$ = Chi-Square test.

### Group-wise comparisons of lipid hair compounds

To compare ECB or related lipid levels in hair between the two groups, Mann-Whitney *U* tests were conducted. Individuals in the MDD group showed statistically significant differences or trends toward higher levels of the following compounds: PI 38:6 (*U* = 39, *p* = .009, *r* = .48), PI O-38:7 (*U* = 38.9, *p* = .004, *r* = .53), MGDG-O 16:3 (*U* = 29.9, *p* = .010, *r* = .51), PG 12:0 (*U* = 42, *p* = .017, *r* = .46), and LPI 13:0 (*U* = 60, *p* = .043, *r* = .37). Significantly lower levels were observed in the MDD group for PI 37:4 (*U* = 20, *p* = .031, *r* = .48) and PI 24:0 (*U* = 19, *p* < .001, *r* = .67). After Bonferroni-Holm p-value adjustment, differences remained statistically significant for PI 38:6 (*p*_adj_ = .045), PI 38:7 (*p*_adj_ = .024), MGDG-O 16:3 (*p*_adj_ = .045), and PI 24:0 (*p*_adj_ < .001). Trends toward significance remained for PI 37:4 (*p*_adj_ = .062), PG 12:0 (*p*_adj_ = .051), and LPI 13:0 (*p*_adj_ = .062). All group comparisons are summarized in Figure 1.

**Figure 1:**
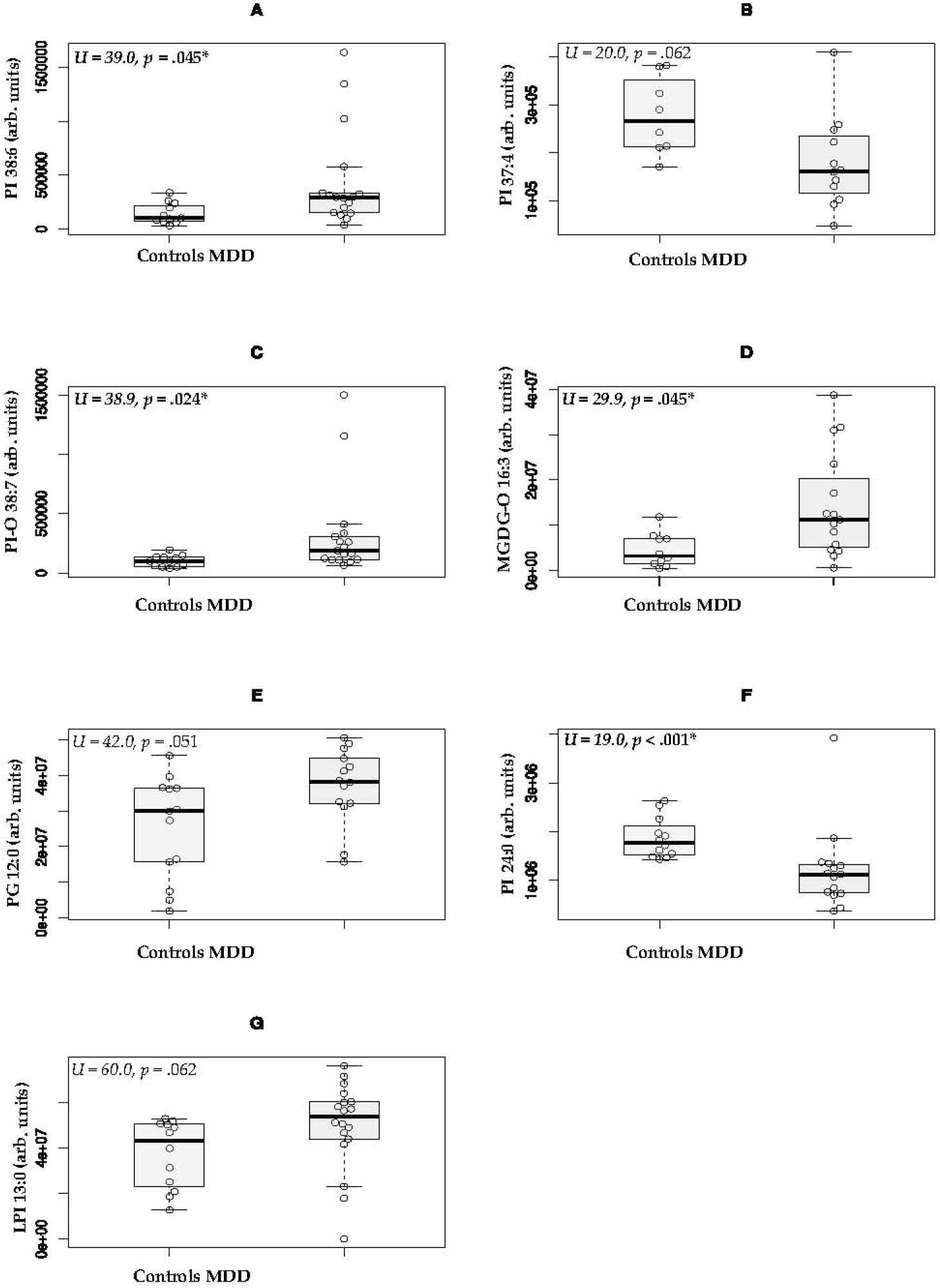
Differences in hair content of lipid compounds between groups. *Notes:* Arb. Units = Arbitrary units; values reported as abundance score; *U* = Mann-Whitney *U* test statistic; *p* = *p*-value after correction; * = indicating statistical significance at α = .05. Group comparisons of hair content of PI 38:6 (A), PI 37:4 (B), PI 0-38:7 (C), MDGD-0 16:3 (D), PG 12:0 (E), PI24:0 (F) and LPI 13:0(G) between individuals with Major Depressive Disorder (MDD) and non-MDD controls. In the MDD group, levels of PI 38:6, PI O-38:7, MGDG-O 16:3, PG 12:0, LPI 13:0 were generally higher, while levels of PI 37:4 and PI 24:0 were lower compared to the control group.

### Regression analyses depending on the severity of depressive symptoms

For lipid compounds that showed significant differences between the MDD and control groups, separate linear regression models were fit for the total cohort, using the sum scores of both the BDI (self-report) and the MADRS (interview) as dependent variables. The BDI-dependent models are summarized in Figure 2. The compound PI 24:0 showed a significant effect (*F*1, 25 = 9.98, *p* = .004, *R*² = .29, *B* = −9.99E-6, 95% CI [-1.60E-5, −3.00E-6]), and four other compounds showed relevant trends in explaining variance in the BDI sum scores. These were LPI 13:0 (*F*1, 28 = 2.59, *p* = .118, *R*² = .09), PI 37:4 (*F*1, 18 = 4.23, *p* = .054, *R*² = .19), PI-O 38:7 (*F*1, 27 = 2.62, *p* = .117, *R*² = .09), and PG 12:0 (*F*1, 25 = 2.64, *p* = .117, *R*² = .10).

**Figure 2:**
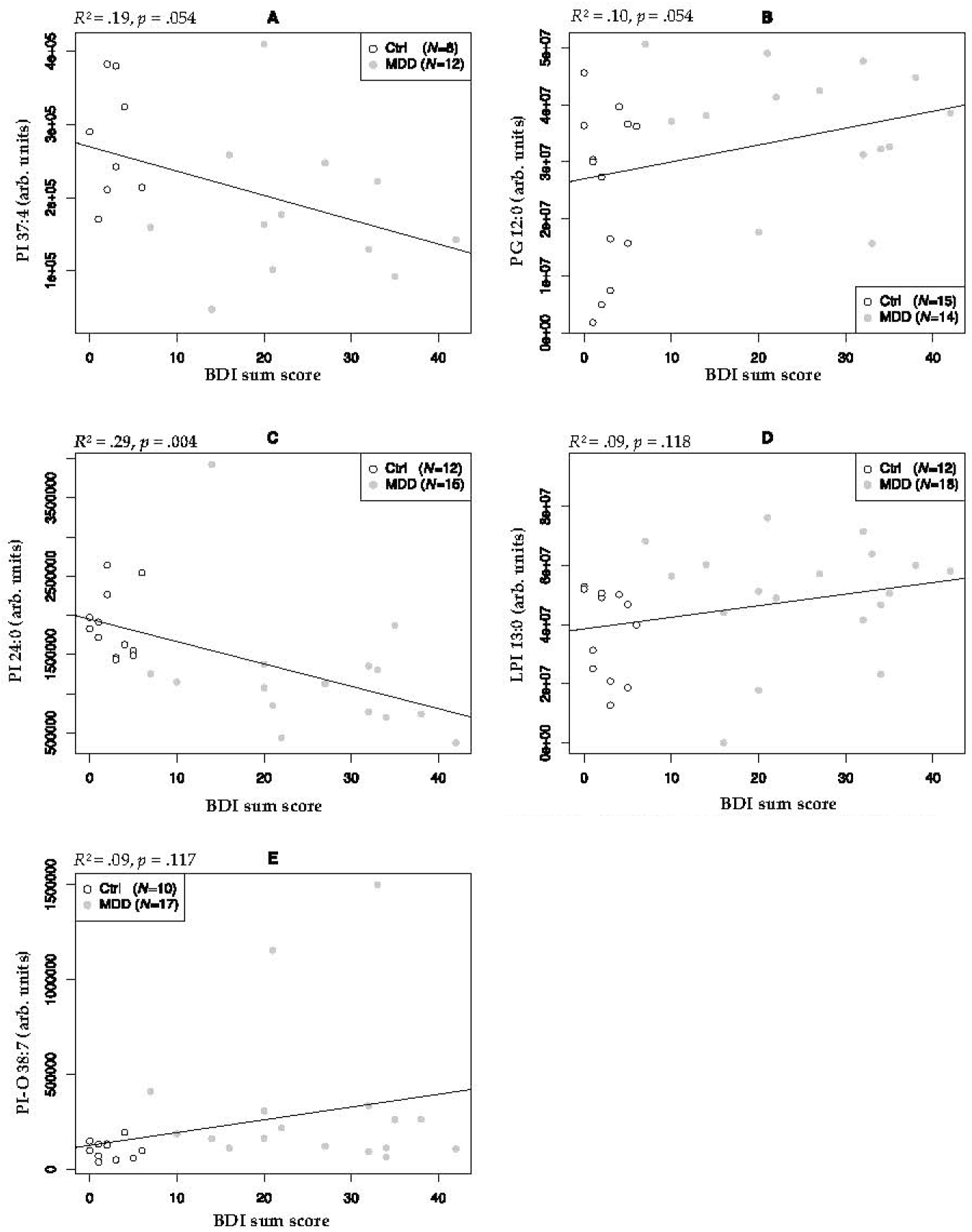
Associations between members of the endocannabinoid family detected in hair samples (3 cm strands) and the clinical severity of depressive symptoms. *Notes:* BDI = *Beck’s Depression Inventory*; arb. units = Abundance score (arbitrary units). Higher depressive symptom severity measured by BDI sum scores was significantly associated with lower levels of PI 24:0, *R²* = .29, and as a trend with lower levels of PI 37:4, *R²* = .19 (A) and higher levels of PG 12:0, *R²* = .10 (B), PI-O 38:7, *R²* = .09, and LPI 13:0, *R²* = .09.

Higher depressive symptom severity (self-report) was associated with lower levels of PI 24:0 and PI 37:4, and higher levels of LPI 13:0, PI-O 38:7, and PG 12:0. Repeated analysis using MADRS scores revealed significant effects for PI 24:0 (*F*1, 25 = 6.92, *p* = .014, *R*² = .22, *B* = −8.46E-6, 95% CI [-1.50E-5, −2.00E-6]) and PI 37:4 (*F*1, 18 = 5.53, *p* = .030, *R*² = .24, *B* = - 6.30E-5, 95% CI [-1.19E-4, −7.00E-6]), with trends toward significance for PG 12:0 (*F*1, 25 = 3.56, *p* = .071, *R*² = .12), LPI 13:0 (*F*1, 28 = 2.64, *p* = .115, *R*² = .09), and PI-O 38:7 (*F*1, 27 = 3.46, *p* = .074, *R*² = .11). As with the BDI-dependent associations, higher MADRS scores were associated with lower levels of PI 24:0 and PI 37:4, and higher levels of LPI 13:0, PI-O 38:7, and PG 12:0. Table 2 presents and compares the regression model values based on the sum scores of BDI and MADRS, respectively.

**Table 2:**
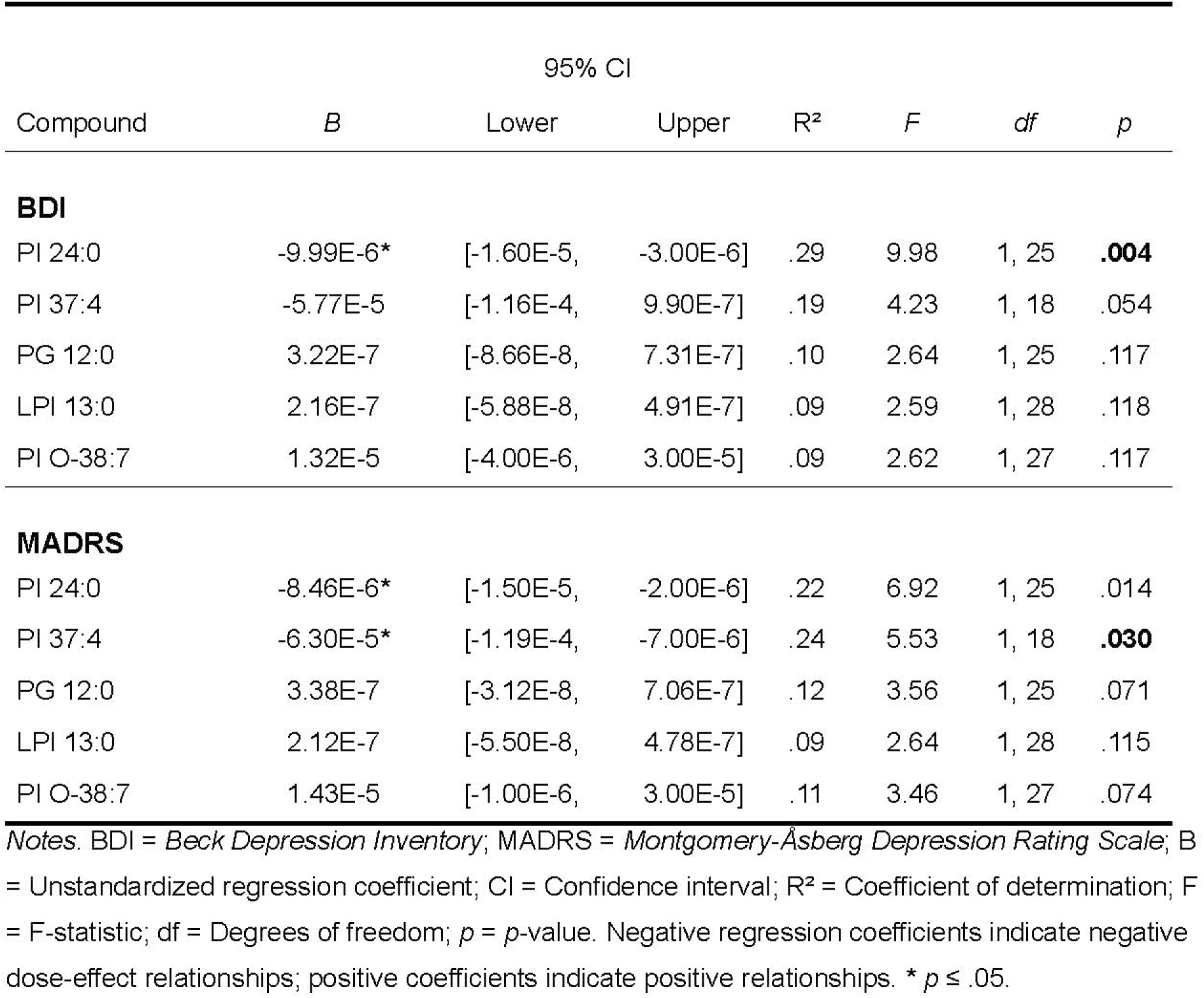
Comparison of regression models reflecting dose-effect relationships depending on BDI and MADRS sum scores.

| Compound | *B* | 95% CI |  | R<sup>2</sup> | *F* | *df* | *p* |
| --- | --- | --- | --- | --- | --- | --- | --- |
|  |  | Lower | Upper |  |  |  |  |
| **BDI** |  |  |  |  |  |  |  |
| PI 24:0 | -9.99E-6* | [-1.60E-5, | -3.00E-6] | .29 | 9.98 | 1, 25 | **.004** |
| PI 37:4 | -5.77E-5 | [-1.16E-4, | 9.90E-7] | .19 | 4.23 | 1, 18 | .054 |
| PG 12:0 | 3.22E-7 | [-8.66E-8, | 7.31E-7] | .10 | 2.64 | 1, 25 | .117 |
| LPI 13:0 | 2.16E-7 | [-5.88E-8, | 4.91E-7] | .09 | 2.59 | 1, 28 | .118 |
| PI O-38:7 | 1.32E-5 | [-4.00E-6, | 3.00E-5] | .09 | 2.62 | 1, 27 | .117 |
| **MADRS** |  |  |  |  |  |  |  |
| PI 24:0 | -8.46E-6* | [-1.50E-5, | -2.00E-6] | .22 | 6.92 | 1, 25 | .014 |
| PI 37:4 | -6.30E-5* | [-1.19E-4, | -7.00E-6] | .24 | 5.53 | 1, 18 | **.030** |
| PG 12:0 | 3.38E-7 | [-3.12E-8, | 7.06E-7] | .12 | 3.56 | 1, 25 | .071 |
| LPI 13:0 | 2.12E-7 | [-5.50E-8, | 4.78E-7] | .09 | 2.64 | 1, 28 | .115 |
| PI O-38:7 | 1.43E-5 | [-1.00E-6, | 3.00E-5] | .11 | 3.46 | 1, 27 | .074 |

Notes. BDI = Beck Depression Inventory; MADRS = Montgomery-Åsberg Depression Rating Scale; *B* = Unstandardized regression coefficient; CI = Confidence interval; $R^2$ = Coefficient of determination; *F* = F-statistic; *df* = Degrees of freedom; *p* = *p*-value. Negative regression coefficients indicate negative dose-effect relationships; positive coefficients indicate positive relationships. \* $p \leq .05$ .

*2.3 Correlation analysis of ECB levels in hair and mitochondrial bioenergetics in PBMC* Finally, possible correlations between the identified hair lipids and mitochondrial bioenergetics in PBMC were examined. Several lipids correlated with measures of mitochondrial respiratory activity in PBMC (Table 3). Notably, lipids that differed between depressed and non-depressed individuals were closely related to mitochondrial function. For example, *Uncoupled* respiration correlated negatively with PG 12:0 (*r* = -.48, *p* = .022) and MGDG-O 16:3 (*r* = -.49, *p* = .022) - both compounds were higher in the MDD group - and positively with PI 24:0 (*r* = .47, *p* = .027) – which was lower in the MDD group. Following the same pattern, ATP turnover-related respiration was positively associated with PI 24:0 (*r* = .33, *p* = .030) and negatively with MGDG-O 16:3 (*r* = -.45, *p* = .033).

**Table 3:**
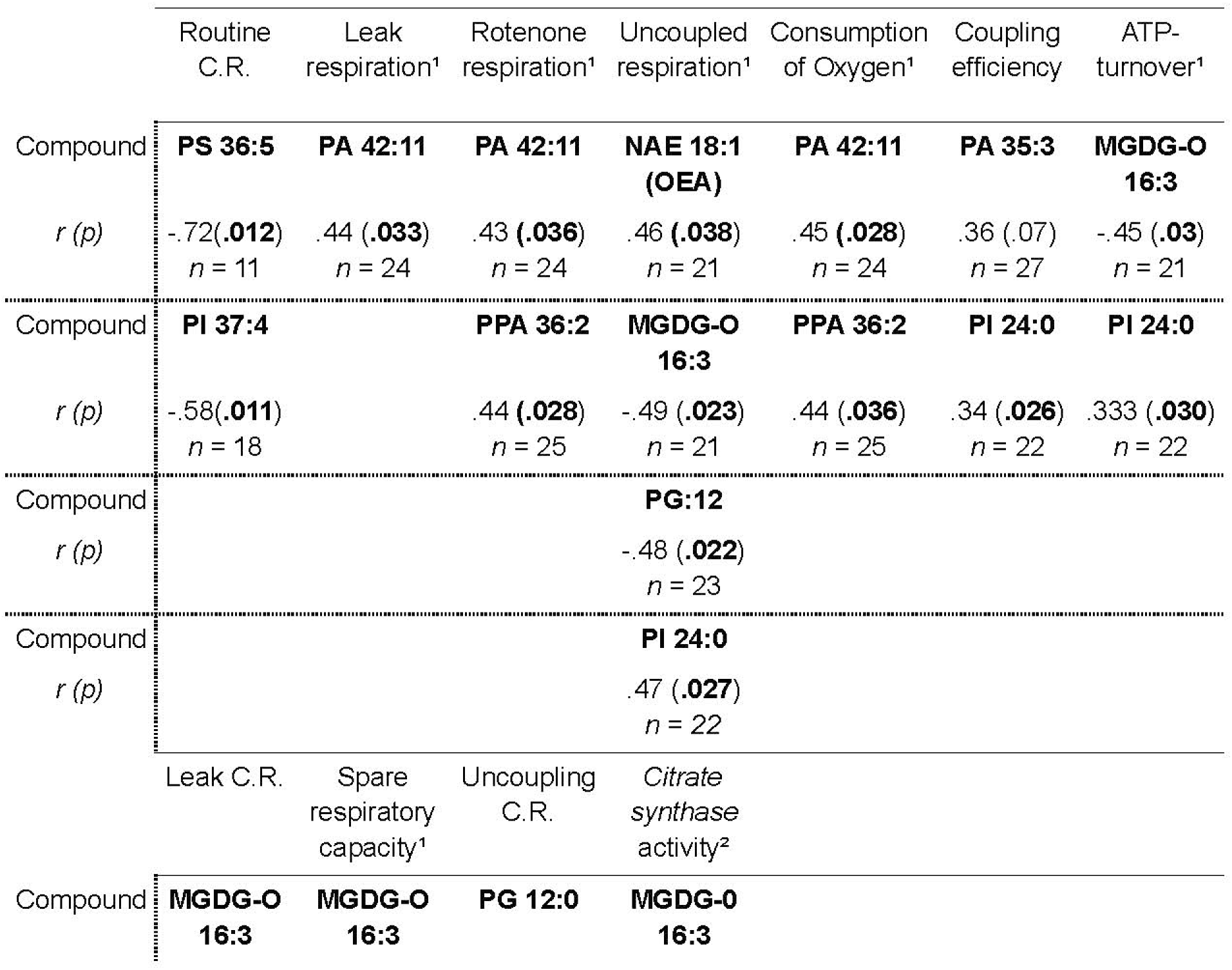
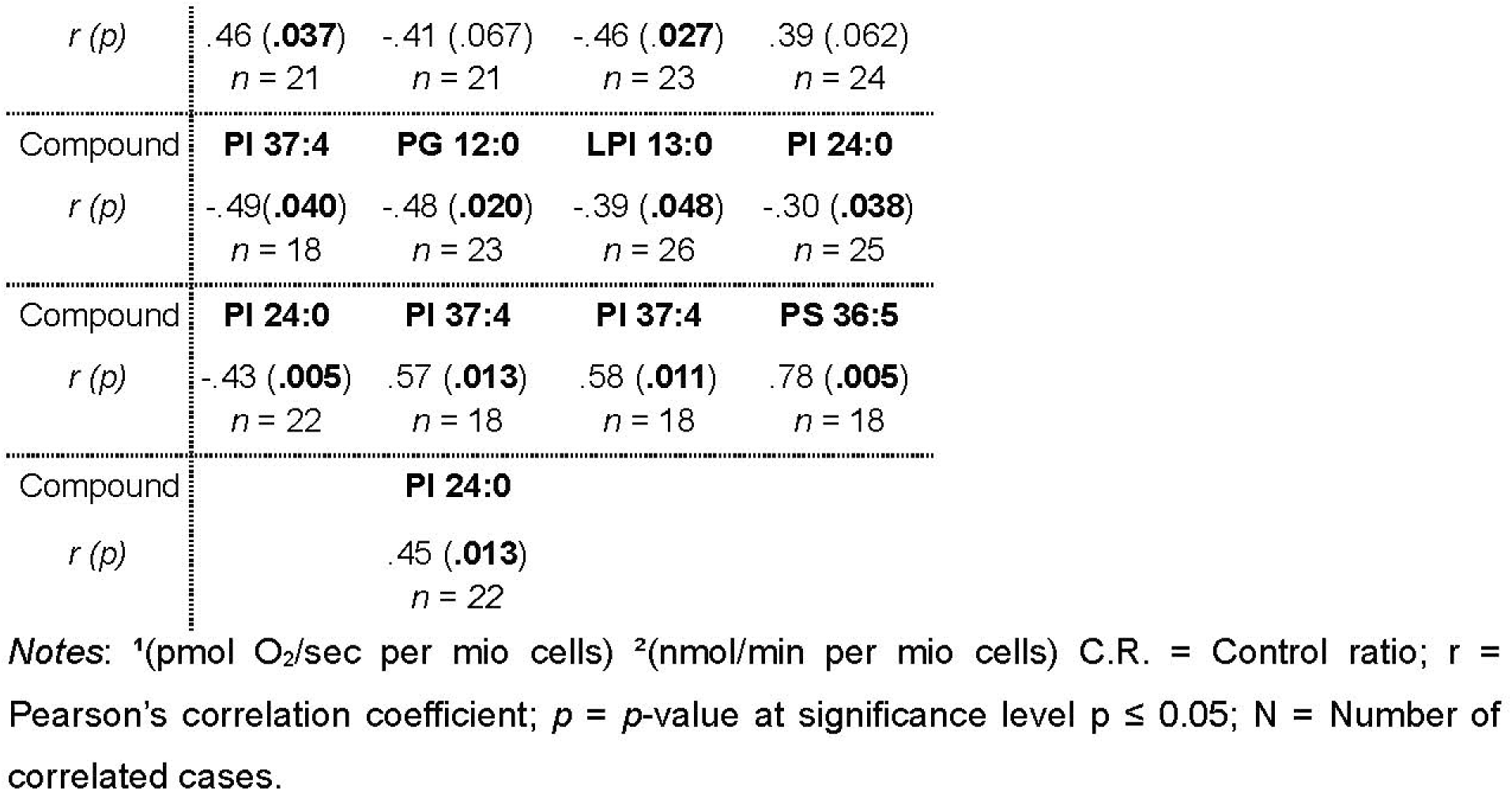
Significant correlations between measures of mitochondrial respiratory activity in PBMCs and lipid compounds in hair.

|  | Routine<br>C.R. | Leak<br>respiration<sup>1</sup> | Rotenone<br>respiration<sup>1</sup> | Uncoupled<br>respiration<sup>1</sup> | Consumption<br>of Oxygen<sup>1</sup> | Coupling<br>efficiency | ATP-turnover<sup>1</sup> |
| --- | --- | --- | --- | --- | --- | --- | --- |
| Compound | **PS 36:5** | **PA 42:11** | **PA 42:11** | **NAE 18:1**<br>**(OEA)** | **PA 42:11** | **PA 35:3** | **MGDG-O**<br>**16:3** |
| *r* (*p*) | -.72(.012)<br>*n* = 11 | .44 (.033)<br>*n* = 24 | .43 (.036)<br>*n* = 24 | .46 (.038)<br>*n* = 21 | .45 (.028)<br>*n* = 24 | .36 (.07)<br>*n* = 27 | -.45 (.03)<br>*n* = 21 |
| Compound | **PI 37:4** |  | **PPA 36:2** | **MGDG-O**<br>**16:3** | **PPA 36:2** | **PI 24:0** | **PI 24:0** |
| *r* (*p*) | -.58(.011)<br>*n* = 18 |  | .44 (.028)<br>*n* = 25 | -.49 (.023)<br>*n* = 21 | .44 (.036)<br>*n* = 25 | .34 (.026)<br>*n* = 22 | .333 (.030)<br>*n* = 22 |
| Compound |  |  |  | **PG:12** |  |  |  |
| *r* (*p*) |  |  |  | -.48 (.022)<br>*n* = 23 |  |  |  |
| Compound |  |  |  | **PI 24:0** |  |  |  |
| *r* (*p*) |  |  |  | .47 (.027)<br>*n* = 22 |  |  |  |
|  | Leak C.R. | Spare<br>respiratory<br>capacity<sup>1</sup> | Uncoupling<br>C.R. | *Citrate*<br>*synthase*<br>*activity*<sup>2</sup> |  |  |  |
| Compound | **MGDG-O**<br>**16:3** | **MGDG-O**<br>**16:3** | **PG 12:0** | **MGDG-O**<br>**16:3** |  |  |  |

|  |  |  |  |  |
| --- | --- | --- | --- | --- |
| *r* (*p*) | .46 (.037)<br>*n* = 21 | -.41 (.067)<br>*n* = 21 | -.46 (.027)<br>*n* = 23 | .39 (.062)<br>*n* = 24 |
| Compound | **PI 37:4** | **PG 12:0** | **LPI 13:0** | **PI 24:0** |
| *r* (*p*) | -.49(.040)<br>*n* = 18 | -.48 (.020)<br>*n* = 23 | -.39 (.048)<br>*n* = 26 | -.30 (.038)<br>*n* = 25 |
| Compound | **PI 24:0** | **PI 37:4** | **PI 37:4** | **PS 36:5** |
| *r* (*p*) | -.43 (.005)<br>*n* = 22 | .57 (.013)<br>*n* = 18 | .58 (.011)<br>*n* = 18 | .78 (.005)<br>*n* = 18 |
| Compound | **PI 24:0** |  |  |  |
| *r* (*p*) | .45 (.013)<br>*n* = 22 |  |  |  |

Notes: <sup>1</sup>(pmol O<sub>2</sub>/sec per mio cells) <sup>2</sup>(nmol/min per mio cells) C.R. = Control ratio; *r* = Pearson's correlation coefficient; *p* = *p*-value at significance level *p* ≤ 0.05; *N* = Number of correlated cases.

The *Leak control ratio* (*Leak* respiration: *Uncoupled* respiration) was associated with higher levels of MGDG-O 16:3 (*r* = .46, *p* = .037) and lower levels of PI 37:4 (*r* = -.49, *p* = .040) and PI 24:0 (*r* = -.43, *p* = .005). *Spare respiratory capacity* (*Uncoupled* – *Routine* respiration) was associated with lower levels of MGDG-O 16:3 (*r* = -.41, *p* = .067) and PG 12:0 (*r* = -.48, *p* = .020), and with higher levels of PI 37:4 (*r* = .57, *p* = .013) and PI 24:0 (*r* = .45, *p* = .013). Furthermore, *Net Routine* (ATP turnover-related respiration: *Uncoupled* respiration) correlated negatively with PG 12:0 (*r* = -.46, *p* = .027) and positively with PI 37:4 (*r* = .58, *p* = .011). Finally, *citrate synthase* activity, spectrophotometrically measured to assess mitochondrial density in PBMCs, was positively associated with MGDG-O 16:3 (*r* = .39, *p* = .062) and negatively with PI 24:0 (*r* = -.30, *p* = .038). An exemplary visual representation of these correlations is provided in Appendix C.

## Discussion

Depression remains a psychiatric disorder whose pathophysiology is not yet fully understood. Advances in Omics sciences and mitochondrial bioenergetics have provided new starting points for identifying biochemical fingerprints and functional signatures of bioenergetic impairment in MDD, toward the development of clinically applicable biomarkers. Here, we combined these conceptual and methodological approaches and first found that several lipids differed significantly in 3cm hair segments from patients with MDD compared with MDD-free controls. Moreover, these lipids were strongly correlated with one another, suggesting that their association with MDD may reflect shared pathways, mechanisms, or interdependencies. Of the seven lipids associated with MDD, five were GPs from the glycerophosphoinositol subgroup (PI 38:6, PI O-38:7, LPI 13:0, PI 37:4, and PI 24:0), one was from the glycerophosphoglycerol subgroup (PG 12:0), and one was a GL from the glycosyldiradylglycerol subgroup (MGDG O-16:3). According to the literature, PIs, along with other GP and GL species, have served as biomarkers for a variety of somatic health conditions, including obesity, diabetes, myopathy, and pancreatic cancer (Hu et al., 2009), as well as mental health conditions such as depression and PTSD (Karabatsiakis et al., 2015; Walther et al., 2018). In the current study, PG 12:0 from the glycerophosphoglycerol subgroup showed a statistically significant dose-response relationship with the severity of depressive symptomatology, while four PIs showed a trend toward significance (LPI 13:0, PI O-38:7, PI 37:4, and PI 24:0). These findings support the idea that lipid compounds in hair might serve as functional biomarkers for depressive disorders. As demonstrated in our previously published data, the presence and severity of depressive symptoms in this cohort were associated with reduced mitochondrial activity (mitochondrial bioenergetics) and changes in mitochondrial density in PBMC (Karabatsiakis et al., 2014). Importantly, analysis of mitochondrial function in PBMC requires invasive blood collection and is technically and methodologically challenging, limiting its applicability as a routine clinical parameter. This limitation is further underscored by the infrastructure requirements for measuring larger sample sizes.

Although it is crucial to understand the role of mitochondrial function in preventive (mitochondrial health index), predictive (bioenergetic health index), and more personalized measures (response to clinical interventions) in MDD, the prerequisites for measuring mitochondrial function in the lab require surrogate markers that are easier to collect, handle, and measure. Hair samples are now considered a very promising candidate for this approach. According to our first observation reported here, lipidomic changes in hair samples seem to reflect alterations in mitochondrial function within immune cells in the context of MDD. The ECS-related lipids identified with this approach can be separated into two groups: The first group of compounds with significantly higher levels in the MDD group (Elevated When Depressed – EWD): PI 38:6; PI-O 38:7; MGDG-O 16:3; PG 12:0; and LPI 13:0; and the second group of those with significantly lower levels in the MDD group (Reduced When Depressed - RWD): PI 24:0 and PI 37:4. As shown in Table 3, lipids in the EWD group correlated negatively with parameters representing the overall efficiency and capacity of mitochondrial ATP production and positively with parameters that might indicate a stressed state of reduced mitochondrial efficiency. The negative associations of these lipids with mitochondrial measures suggest a specific alteration in mitochondrial function, in which both the capacity for ATP production (reflected in ATP turnover-related respiration) and the process of uncoupled respiration (typically involving mechanisms that dissipate the proton gradient without producing ATP) are reduced. Simultaneously, the lipids of the EWD group are associated with a higher *Leak control ratio*, possibly indicating that the remaining respiratory activity is more tightly coupled to ATP synthesis than to proton leak. This pattern might reflect several underlying mitochondrial alterations, such as reduced overall mitochondrial activity or enhanced efficiency under stress. In response to stress or damage, mitochondria might increase coupling efficiency to reduce ROS production, which typically rises during uncoupled or inefficient electron transport. In this context, a higher *Leak control ratio* might reflect a protective mechanism that reduces oxidative stress. Theoretically, lower overall mitochondrial respiration rates alongside a higher *Leak control ratio* could also result from a shift in metabolic strategy from oxidative phosphorylation (OXPHOS) toward glycolysis. So far, this mechanism, known as the Warburg effect (Menendez et al., 2013), has been primarily studied in cancer cells. Because mitochondrial bioenergetics is a relatively new area in psychiatric conditions, all available metabolic mechanisms should be considered when interpreting results. In addition to apparently promoting an overall stressed mitochondrial state, lipids in the EWD group were negatively associated with spare respiratory capacity (*Uncoupled* respiration – *Routine* respiration). This could reflect the mitochondria’s ability to respond to increased energy demands by upregulating ATP production. Downregulation of these parameters might reflect reduced flexibility in energy production, either because mitochondria are already operating at an elevated level, comparable to their uncoupled state (due to innate uncoupling mechanisms or pathological conditions), or because they have limited capacity to increase respiration in response to uncoupling. This could indicate mitochondrial perturbation or an adaptation to reduce ROS production. Conversely, lipids in the RWD group showed the opposite associations with mitochondrial parameters. They were positively correlated with measures of overall mitochondrial efficiency and capacity, including uncoupled respiration, ATP turnover-related respiration, and spare respiratory capacity, and, accordingly, negatively correlated with the leak control ratio. Additionally, PI 24:0, a lipid in the RWD group, was negatively correlated with *citrate synthase* activity, a robust measure of mitochondrial density and biogenesis. In this context, increased mitochondrial biogenesis is likely an adaptation to stress caused by reduced metabolic efficiency.

Taken together, these data support the hypothesis that changes in mitochondrial respiratory function in individuals with MDD (increased mitochondrial biogenesis and decreased mitochondrial bioenergetics) are related to alterations in lipid systems. The current data provide the first empirical evidence that measurable alterations in hair phospholipid composition are linked to and may reflect mitochondrial function in stress-related conditions such as MDD. In conclusion, alterations in lipid systems, particularly within the ECS, warrant further investigation into mitochondrial function and other mental health conditions. By identifying lipid species associated with MDD and elucidating the pathophysiology of mitochondrial bioenergetics, this study contributes to the landscape of the lipidome and/or endocannabinoidome relevant to MDD pathology. This may also be relevant in the future, as ECB synthesis requires phospholipids extracted from cell membrane lipid bilayers. The aspect of “membrane health” has become of interest in cellular stress research (Husted and Bouzinova, 2016; Maccarrone et al., 2011; Schneider et al., 2017). Since the demand for ECB production is closely linked to psychological and/or molecular stress, such events regulate and alter the dynamic composition of cell membranes, thereby affecting their microphysical properties and functionality in synaptic transmission. Fluidity properties of the cell membrane are considered to play a crucial role in the functioning and plasticity of neuronal systems, while dysregulations of lipid systems and subsequent alterations of the membrane have been suggested to be part of the etiopathology of mood disorders by altering neurotransmitter-based signaling and thereby compromising the membrane health of cells and tissues (Maccarrone et al., 2011; Schneider et al., 2017; Walther et al., 2018).

### Limitations

Despite these initially promising findings, several limitations must be considered. First, only age-progressed, postmenopausal females were studied, limiting the generalizability of the findings to both sexes and to younger individuals. Females with depression were receiving antidepressant medication, which might have contributed to group differences. Furthermore, the MDD and control groups differed significantly in BMI and smoking status, both of which are known to influence endocannabinoid signaling and lipid metabolism. Because the small sample size precluded the inclusion of these variables as covariates, their potential contribution to the observed group differences cannot be ruled out and should be addressed in future studies with larger cohorts. Additionally, long-term dietary patterns are a meaningful, unaddressed confound in hair-based lipidomics research, given that food intake is a primary determinant of lipid availability and endocannabinoid tone. Although hair analysis smooths short-term fluctuations, dietary habits were not systematically assessed in the present study and may have contributed to interindividual variability in the observed lipid levels. Moreover, the identified correlations and associations do not clarify the distinct regulatory mechanisms underlying the observations in this study, and the findings must be replicated in larger samples to demonstrate the robustness of our results. In addition to cross-sectional studies, future interventional studies should investigate the reversibility of the biological alterations found in MDD.

From a methodological perspective, the lipid values were obtained using an untargeted discovery approach and therefore do not reflect quantitative levels. The compound candidates identified here should be tested using a targeted approach with the respective standards, which were not commercially available at the time of laboratory assessments. Nevertheless, the ECB biomarker candidates proposed here might serve as a useful tool for assessing physiological burden and monitoring symptom normalization following clinical interventions. As translationally oriented research on the effects of ECB signaling on mitochondrial bioenergetics, especially in the context of mental disorders, has just started, it can be assumed that this link might reveal other central aspects of the ECS in the pathophysiological processes associated with MDD.

## Statements

## Acknowledgment

We thank all participants for their support and compliance throughout the study.

## Statement of Ethics

### Institutional Review Board Statement

The study was conducted in accordance with the Declaration of Helsinki and approved by the Ethics Committee of the Hanover Medical School in Hanover, Germany (protocol code 5747, initially approved on 2011-01-11).

### Informed Consent Statement

Informed consent was obtained from all subjects involved in the study.

## Author Contributions

Conceptualization and realization of the project were conducted by Alexander Karabatsiakis (AK) with support from Detlef E. Dietrich (DED) as the principal investigator (PI) of the project, located at the AMEOS Clinic for Psychiatry and Psychotherapy Hildesheim. In collaboration with Juan Salinas Manrique (JSM), who was involved in clinical assessments and in collecting hair samples from depressed and non-depressed individuals, AK generated the hair sample cohort, processed and analyzed the samples, generated the biological data, and performed the statistical processing of the results. KdP supported the data analyses and provided editorial assistance. Leonard Bondy (LB) contributed to the processing of raw data, further elaborated and finalized statistics, and established the interpretation of results together with AK. Thomas Hennessy (TH) and Thomas Stoll (TS) supported the mass spectrometry analyses under the supervision of Michelle M. Hill (MHM). LB generated the manuscript together with AK, and all authors who have read and agreed to the final version of the manuscript provided input.

## Data Availability Statement

Data presented in this manuscript will not be made available to online repositories but can be made available in a pseudonymized form upon request.

## Appendices

## Appendix A List of ECB-related compounds detected by mass spectrometry

|  | Group |  |  |  |
| --- | --- | --- | --- | --- |
|  | Controls |  | MDD |  |
|  | Mean<br>(arb. units) | N | Mean<br>(arb. units) | N |
| *Lipid compounds* |  |  |  |  |
| NAE 20:0 (Eicosanoyl-EA) |  |  | 355045 | 11 |
| NAE 17:0 (Margaroyl-EA) | 340844 | 7 |  |  |
| NAE 18:1 (Oleoyl-EA) | 1800360 | 9 | 2177413 | 15 |
| NAE 15:0 (Pentadecanoyl-EA) | 2092403 | 10 |  |  |
| MGDG-O 16:3 | 4502547 | 10 | 14375103 | 15 |
| PE 40:10 | 6196338 | 10 |  |  |
| LPG 22:1 | 723425 | 11 |  |  |
| PG 12:0 | 25295243 | 13 | 37101803 | 14 |
| PG O-42:7 |  |  | 299458 | 10 |
| PI 24:0 | 1870642 | 12 | 1219368 | 15 |
| LPI 13:0 | 37532509 | 12 | 49784014 | 18 |
| LPI 15:0 | 8484576 | 6 |  |  |
| PI 34:0 | 419432 | 8 |  |  |
| LPI 22:1 | 191386 | 6 |  |  |
| PI O-36:5 | 178780 | 7 |  |  |
| LPS 16:0 | 120432 | 10 |  |  |
| PS 36:5 |  |  | 1749214 | 11 |
| TG 65:9 | 321987 | 7 |  |  |
| MGDG 36:10 |  |  | 181241 | 12 |
| PA 39:1 | 95049 | 10 |  |  |
| PA 35:3 | 283605 | 10 | 333834 | 17 |
| PA 42:11 | 129368 | 12 | 147898 | 16 |
| PA 44:3 | 150331 | 13 | 211565 | 17 |
| PG O-28:0 | 264491 | 12 | 491334 | 15 |
| PG O-33:0 | 271460 | 9 |  |  |
| PG O-36:0 | 367037 | 9 | 293141 | 14 |
| PG O-39:0 | 3567363 | 13 |  |  |
| PI 38:6 | 145976 | 11 | 438571 | 17 |
| PI 37:4 | 276579 | 8 | 179224 | 12 |
| PI 36:4 | 196663 | 9 | 142294 | 13 |
| PI 40:6 |  |  | 240043 | 13 |
| PI O-38:7 | 100434 | 12 | 328091 | 17 |
| PPA 36:2 | 193598 | 13 | 255231 | 16 |

## Appendix B Significant correlations between lipid species.

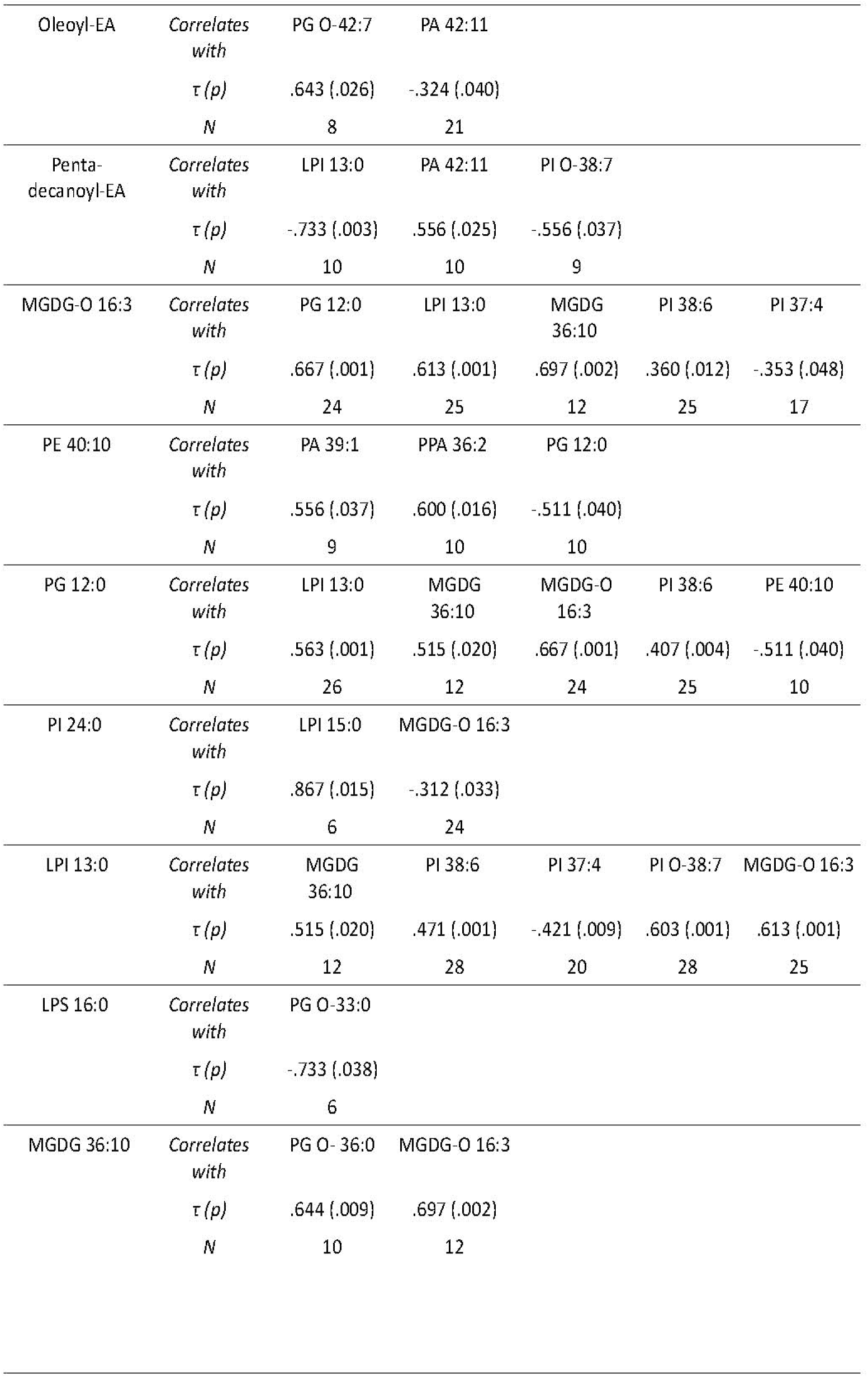
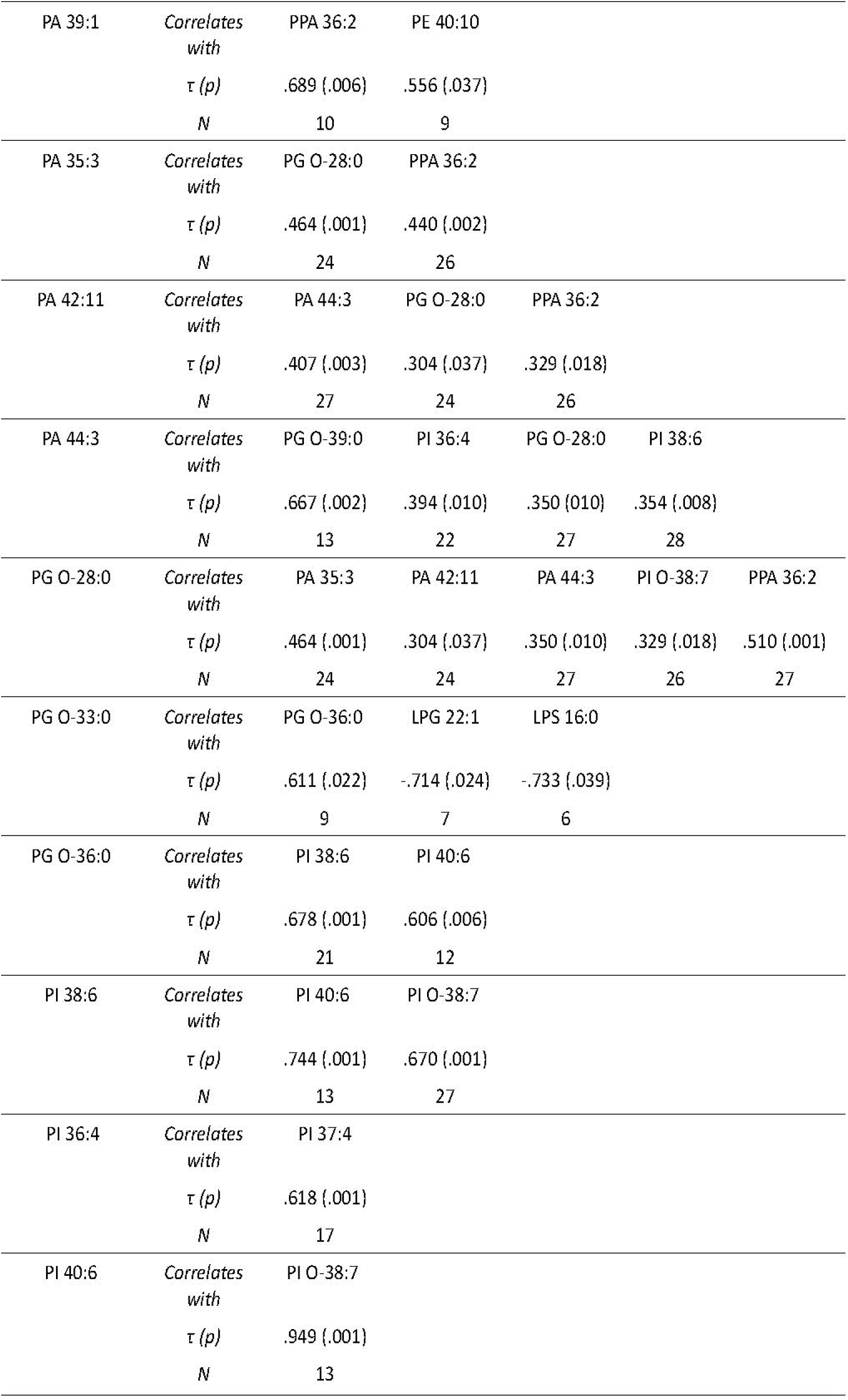
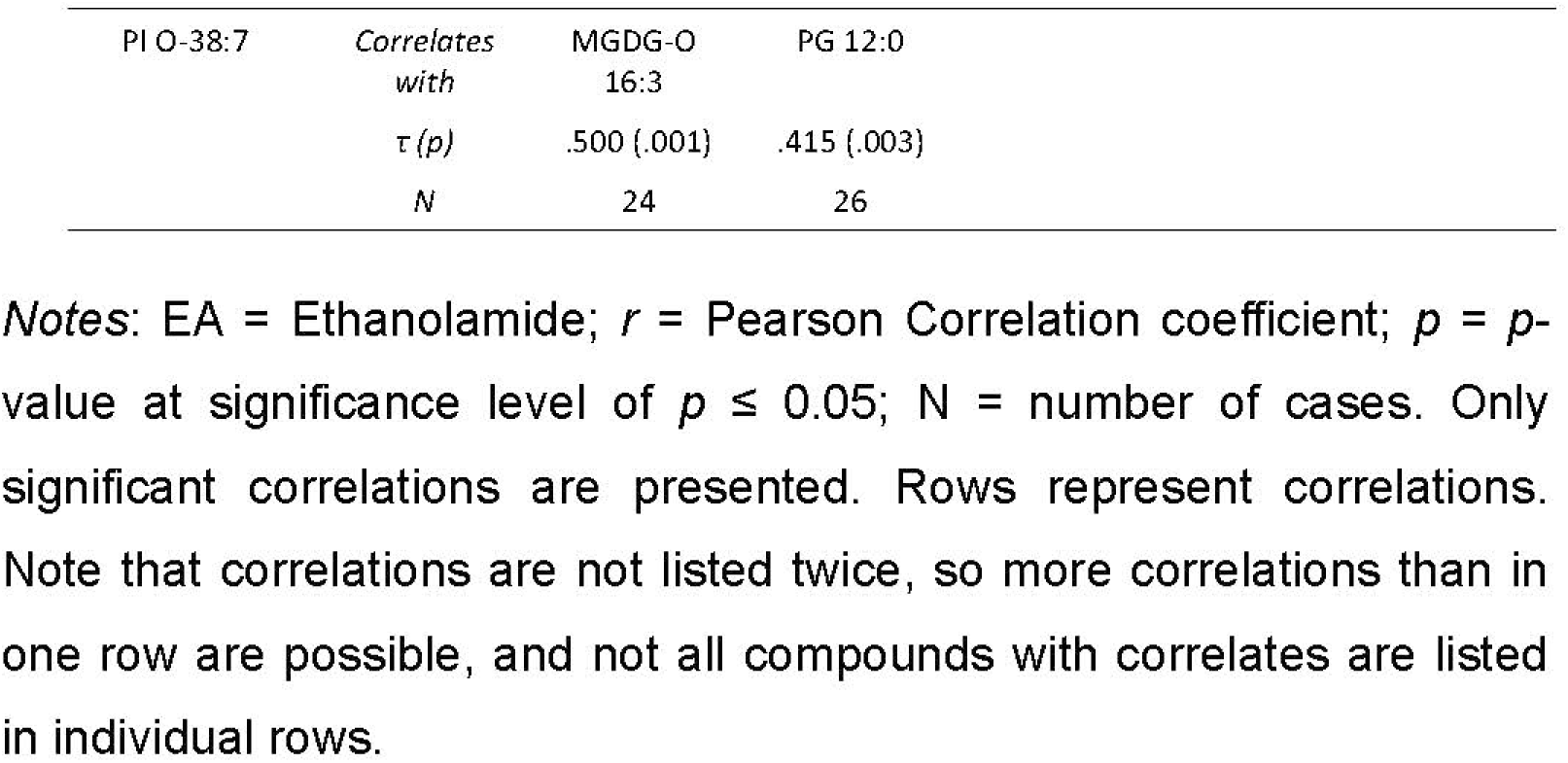

## Appendix C Visual representation of correlations between measures of mitochondrial respiratory activity in PBMC and lipid compounds in hair

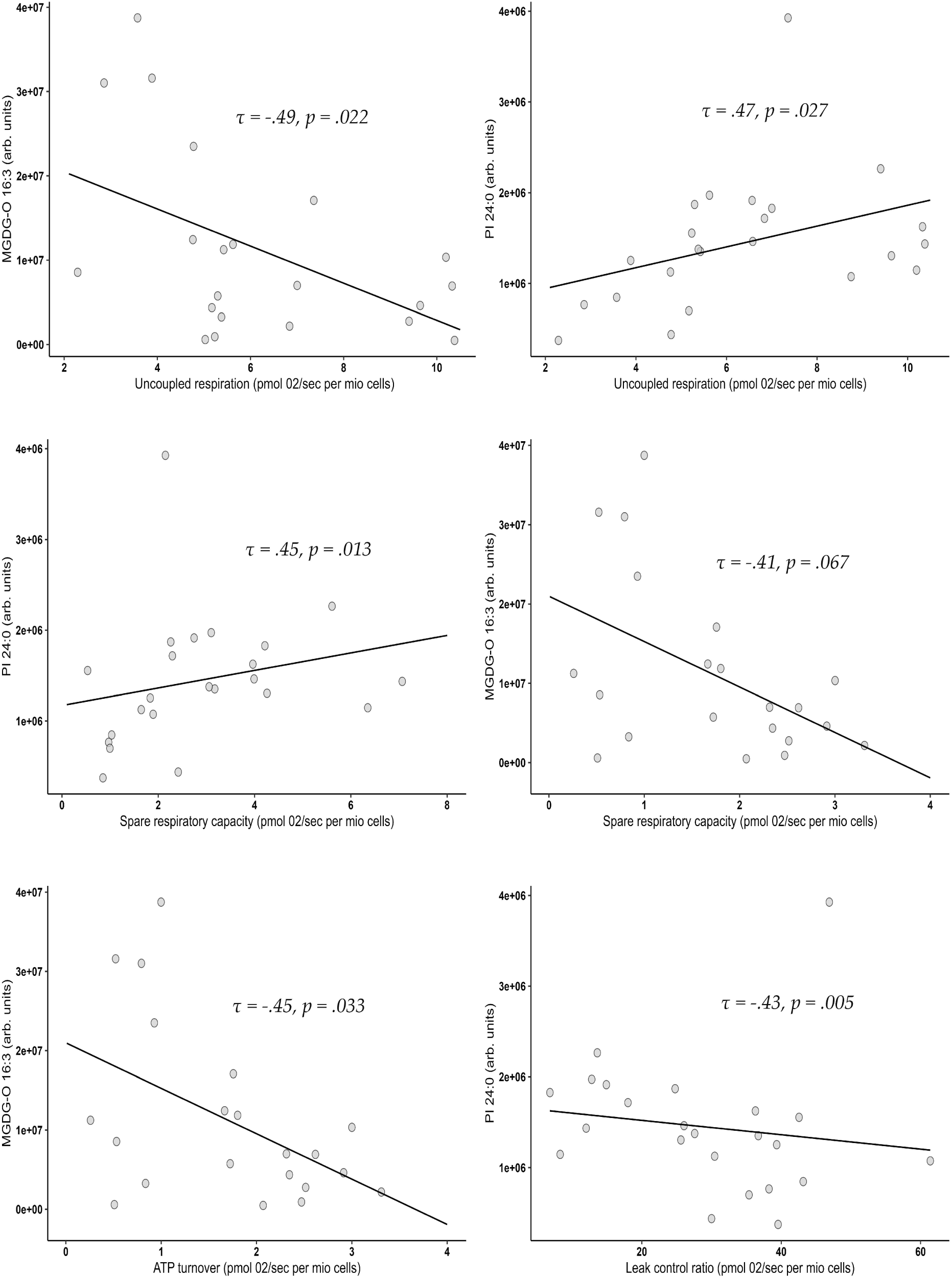

*Notes*: *r* = Pearson Correlation coefficient; *p* = *p*-value at significance level of *p* ≤ 0.05.

